# Endothelium-protective, histone-neutralizing properties of the polyanionic agent defibrotide

**DOI:** 10.1101/2021.02.21.21252160

**Authors:** Hui Shi, Alex A. Gandhi, Stephanie A. Smith, Qiuyu Wang, Diane Chiang, Srilakshmi Yalavarthi, Ramadan A. Ali, Chao Liu, Gautam Sule, Pei-Suen Tsou, Yu Zuo, Yogendra Kanthi, Evan A. Farkash, Jiandie D. Lin, James H. Morrissey, Jason S. Knight

## Abstract

Neutrophil-mediated activation and injury of the endothelium play a role in the pathogenesis of diverse disease states ranging from autoimmunity to cancer to COVID-19. Neutralization of cationic proteins (such as neutrophil extracellular trap/NET-derived histones) with polyanionic compounds has been suggested as a potential strategy for protecting the endothelium from such insults. Here, we report that the FDA-approved polyanionic agent defibrotide (a pleiotropic mixture of oligonucleotides) directly engages histones and thereby blocks their pathological effects on endothelium. *In vitro*, defibrotide counteracted endothelial cell activation and pyroptosis-mediated cell death, whether triggered by purified NETs or recombinant histone H4. *In vivo*, defibrotide stabilized the endothelium and protected against histone-accelerated inferior vena cava thrombosis in mice. Mechanistically, defibrotide demonstrated direct and tight binding to histone H4 as detected by both electrophoretic mobility shift assay and surface plasmon resonance. Taken together, these data provide insights into the potential role of polyanionic compounds in protecting the endothelium from thromboinflammation with potential implications for myriad NET- and histone-accelerated disease states.

## INTRODUCTION

Neutrophils are the most abundant innate effector cells of the human immune system, exerting antimicrobial effects through phagocytosis and degranulation (1). The release of neutrophil extracellular traps (**NETs**)—web-like structures composed of microbicidal cytosolic and granule proteins enmeshed in decondensed chromatin—is a recently described strategy by which neutrophils kill microbes in tissues (2). However, when formed intravascularly, NETs are potentially noxious, trapping red blood cells, activating platelets, and damaging the endothelium—thereby promoting coagulation, vascular occlusion, and thrombosis (3–6). Endothelial activation and injury driven by NETs has been revealed as a key pathogenic step in a variety of disease states including deep vein thrombosis (7), transfusion-related acute lung injury (8), atherosclerosis (9), and lupus (10). High levels of NETs have also been detected in the blood of coronavirus disease 2019 (COVID-19) patients (11), where they likely contribute to the endothelial damage regularly noted on the histopathology of COVID-19 organs (12–15).

NETs present a variety of highly cationic proteins, including histones, HMGB1, calprotectin, cathepsin G, and LL-37, among others. While these proteins contribute to the capture and inactivation of invading microorganisms, they may also be cytotoxic to host tissues (16). In particular, NET-derived histones account for ∼70% of NET-associated proteins (16) and have been associated with endothelial damage and multiple organ dysfunction in acute states such as sepsis (17), acute pancreatitis (18), acute respiratory distress syndrome (19), and severe trauma (20). High levels of circulating histones (up to 250 μg/ml after trauma) (20) activate and damage endothelial cells via pore formation (21, 22), engagement of innate sensors such as Toll-like receptors (23–25) and the NLRP3 inflammasome (26, 27), and forced release of von Willebrand factor (vWF) (28). On balance, the result is a hypercoagulable state and an increased risk of vascular events including thrombosis. Targeting histones by neutralizing their cationic nature with polyanions has been suggested as an approach to combatting various NET- and histone-associated diseases (29, 30).

Defibrotide is a pleotropic mixture of oligonucleotides (90% single-stranded phosphodiester oligonucleotides and 10% double-stranded) that is derived from porcine intestinal mucosal DNA and which has antithrombotic, fibrinolytic, and anti-inflammatory activities (31, 32). Defibrotide was initially approved for the treatment of thrombophlebitis and as prophylaxis for deep vein thrombosis in Italy (33, 34) (although these approvals are no longer active). Subsequently, it was granted an orphan drug designation by European and American regulatory agencies for the treatment of serious hepatic veno-occlusive disease (VOD) after hematopoietic cell transplantation (Europe) or VOD with renal and/or pulmonary dysfunction post-transplant (United States) (34). Although defibrotide’s mechanisms of action remain incompletely understood, there is evidence that it protects endothelium, modulates platelet activation, potentiates fibrinolysis, decreases thrombin generation and activity, and reduces circulating levels of plasminogen activator inhibitor type 1 (35–39). Defibrotide has also been demonstrated to associate with cationic proteins, for example collagen I (40).

Here, we hypothesized that the polyanionic properties of defibrotide might mitigate activation of and damage to the endothelium by NETs and especially NET-derived cationic proteins. In pursuit of this possibility, we characterized defibrotide’s endothelium-protective properties both *in vitro* and in a mouse model of venous thrombosis.

## RESULTS

### Defibrotide inhibits the activation of cultured endothelial cells by NETs

Human umbilical vein endothelial cells (HUVECs) were cultured with human neutrophil-derived NETs in the presence or absence of defibrotide. Gene transcripts associated with the expression of cell adhesion molecules E-selectin, ICAM-1, and VCAM-1 were then quantified. In all cases, expression was markedly increased by purified NETs, whether those NETs were originally triggered by PMA (**Figure 1A-C**) or calcium ionophore (**Supplementary Figure 1**). These NET-mediated increases were consistently restrained in the presence of defibrotide (**Figure 1A-C, Supplementary Figure 1**). Beyond gene expression, we also confirmed that purified NETs increased surface protein expression of E-selectin, ICAM-1, and VCAM-1 via an in-cell ELISA assay, and that these increases could be mitigated by defibrotide (**Figure 1D-F**). We reasoned that if these expression differences were functionally meaningful, then adhesion of neutrophils to the HUVEC monolayer should track in similar fashion (increased by NETs and decreased by defibrotide). As predicted, calcein-AM-labeled human neutrophils adhered more strongly to NET-activated HUVECs, an effect that was reduced in the presence of defibrotide (**Figure 1G**). Beyond surface adhesion molecules, previous work has also suggested that NETs upregulate expression of tissue factor (TF) by endothelial cells, thereby contributing to the prothrombotic state (41). Here we found that TF was upregulated by NETs whether measured by gene expression (**Figure 1H**) or enzymatic activity (**Supplementary Figure 2**); in both contexts, NET-mediated increases were significantly reduced by defibrotide (**Figure 1H, Supplementary Figure 2**). Finally, we examined the effect of defibrotide on NET-regulated permeability of HUVEC monolayers. Indeed, NETs increased permeability across HUVEC monolayers in as little as one hour, whereas the addition of defibrotide reduced this NET-mediated increase (**Figure 1I**). Taken together, these data support the basic premise of the study, namely that defibrotide can neutralize the activation and permeability of endothelial cells by NETs.

**Figure 1:**
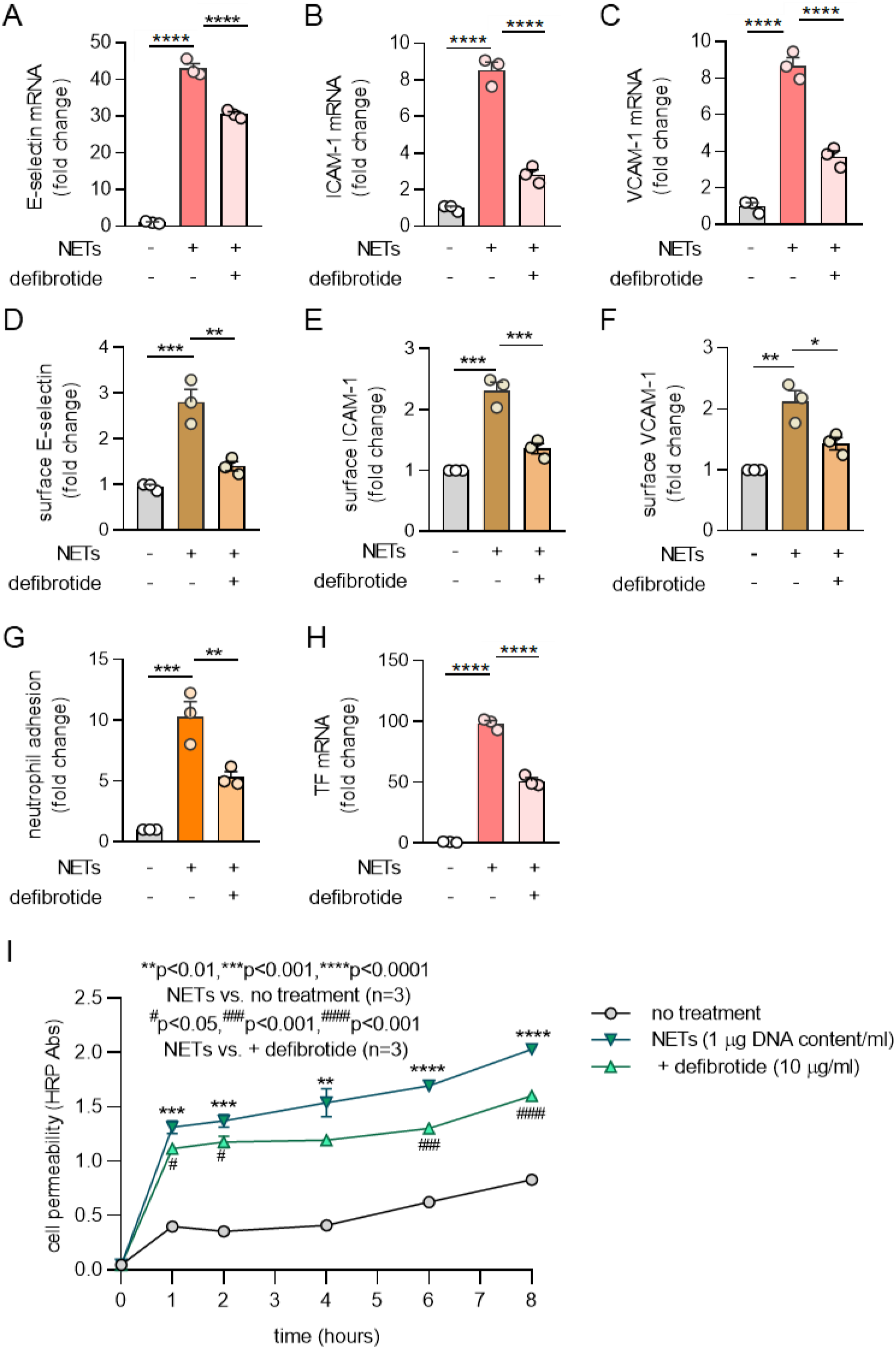
Defibrotide inhibits the activation and permeability of cultured HUVECs by NETs. **A-C,** HUVECs were pretreated with defibrotide (10 μg/ml) for 30 minutes, followed by isolated NETs (1 μg DNA content/ml) for 4 hours. E-selectin (A), ICAM-1 (B), and VCAM-1 (C) mRNA levels were determined by qPCR. Mean ± standard deviation is presented for one representative experiment out of three independent experiments, all with similar results; ****p<0.0001 by one-way ANOVA corrected by Dunnett’s test. **D-F,** HUVECs were pretreated with defibrotide (10 μg/ml) for 30 minutes, followed by the addition of NETs for 6 hours. Surface expression of E-selectin (D), ICAM-1 (E), and VCAM-1 (F) were then detected by in-cell ELISA. **G,** HUVEC monolayers were pretreated with defibrotide (10 μg/ml) for 30 minutes, followed by NETs (1 μg DNA content/ml) for 4 hours. Calcein-AM-labeled neutrophils were then added as described in Methods. Mean ± standard deviation is presented for n=3 independent experiments; **p < 0.01 and ***p < 0.001 by one-way ANOVA corrected by Dunnett’s test. **H,** HUVECs were treated as for panels A-C. Tissue factor mRNA levels were detected at 4 hours. Mean ± standard deviation is presented for n=3 independent experiments; ****p<0.0001 as compared by one-way ANOVA corrected by Dunnett’s test. **I,** HUVECs were treated as for panels A-C. Cell permeability was assessed by measuring horseradish peroxidase (HRP) movement through EC monolayers in a Transwell system as described in Methods. Mean ± standard deviation is presented for one representative experiment out of three independent experiments, all with similar results; **p<0.01, ***p < 0.001 and ****p<0.0001 by two-way ANOVA corrected by Tukey’s test. ^#^p<0.05, ^###^p<0.001, and ^####^p<0.0001 by two-way ANOVA corrected by Tukey’s test.

### Transcriptome profiling confirms a NET-induced proinflammatory signature in endothelial cells, which can be mitigated by defibrotide

The above data demonstrate activation of endothelial cells by NETs in the context of selected genes associated with cell-cell interactions and coagulation. To more broadly understand the pathways associated with endothelial cell activation, we performed RNA sequencing of HUVECs exposed to vehicle, NETs, or NETs + defibrotide. We identified 440 differentially expressed genes (300 upregulated) in HUVECs upon NET stimulation as compared with vehicle. Conversely, there were 229 differentially expressed genes (192 downregulated) when the NETs + defibrotide group was compared to NETs alone. The top upregulated genes are displayed in **Figure 2A**. Functional gene network analysis of upregulated genes in NET-stimulated HUVECs revealed an inflammatory signature highlighted by meta groups such as the TNF signaling pathway, NF-ĸB signaling pathway, and MAPK signaling pathway (**Figure 2B**). Notably, the same pathways that were upregulated by NETs were likely to be downregulated by defibrotide (**Figure 2C**). Taken together, these data confirm the ability of NETs to activate endothelial cells, and demonstrate the ability of defibrotide to reverse those effects.

**Figure 2:**
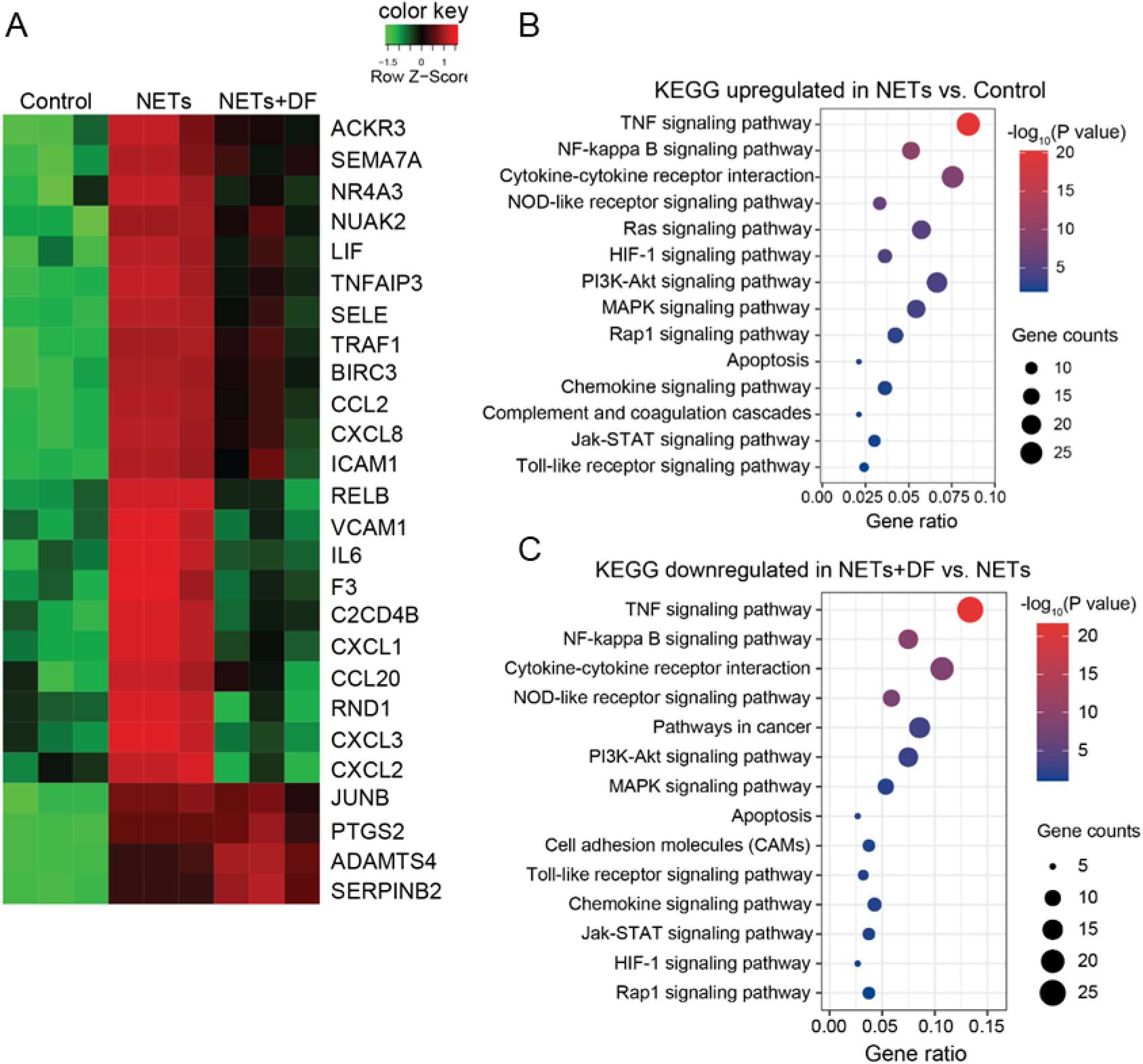
Transcriptome profiling of HUVECs in response to NETs ± defibrotide. **A,** HUVECs were treated with vehicle (PBS), NETs (1 μg DNA content/ml), or NETs+defibrotide (10 μg/ml) for 4 hours (n=3 per group). RNA sequencing was performed. K-means clustering of differentially expressed genes is presented as a heat map. **B,** Bubble plot of upregulated biological processes in the NETs group as compared with the vehicle group. Color-coding is based on p value and bubble size is based on the number of genes in each pathway. **C,** Bubble plot of downregulated biological processes in the NETs group as compared with the NETs+defibrotide group. DF=defibrotide.

### Blocking histone H4 counteracts HUVEC activation by NETs

As discussed above, part of the original hypothesis was that the polyanionic nature of defibrotide might make it especially effective at neutralizing NET-derived cationic proteins such as histones (2), which are important mediators of inflammation, tissue injury, and organ dysfunction in the extracellular space (42, 43). To begin to address this, we asked whether a histone-neutralizing antibody might be effective in our system. Indeed, an anti-histone H4 antibody counteracted the upregulation of HUVEC E-selectin (**Figure 3A**), ICAM-1 (**Figure 3B**), VCAM-1 (**Figure 3C**), and TF (**Figure 3D**) by NETs.

**Figure 3:**
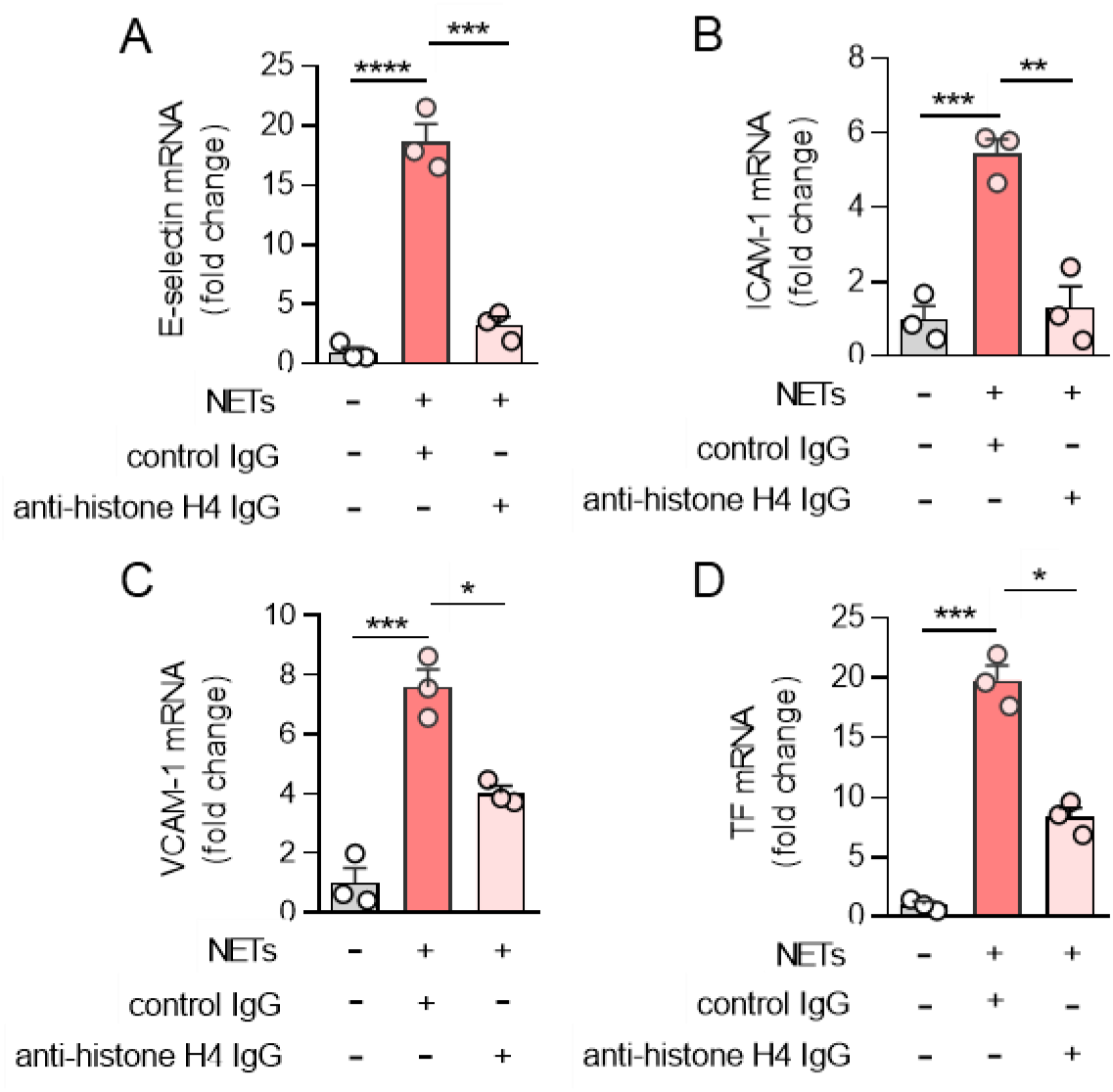
NET-derived histone H4 induces HUVEC activation. **A-D,** NETs (1 μg DNA content/ml) were incubated with antibodies to histone H4 (100 ng/ml) for 1h, and then added to HUVECs for 4 hours. E-selectin (A), ICAM-1 (B), VCAM-1 (C) and tissue factor (TF) mRNA levels were determined by qPCR. Mean ± standard deviation is presented for one representative experiment out of three independent experiments, all with similar results; *p<0.05, **p<0.01, ***p<0.001, and ****p<0.0001 one-way ANOVA corrected by Dunnett’s multiple comparison test.

### Defibrotide abolishes endothelial cell activation by extracellular histone H4

We next asked whether defibrotide might directly antagonize the effects of histone H4. As expected, purified histone H4 increased expression of E-selectin (**Figure 4A**), ICAM-1 (**Figure 4B**), and VCAM-1 (**Figure 4C**) by HUVECs, while defibrotide almost completely abolished these effects. Given that citrullinated histones are an important component of NETs, we also treated HUVECs with citrullinated histone H4. We found similar activation as for native histone H4, and again found that the effect was significantly restrained by defibrotide (**Supplementary Figure 3**); native histone H4 was therefore used in all subsequent experiments. Beyond HUVECs, we questioned whether defibrotide could also protect microvascular endothelial cells against histone H4. We treated human dermal microvascular endothelial cells (HMVECs) with histone H4 and defibrotide, and found a similar pattern as for HUVECs. Specifically, gene transcripts for E-selectin, ICAM-1, VCAM-1, and TF were upregulated by histone H4, and then restrained in the additional presence of defibrotide (**Supplementary Figure 4**). Mechanistically, we found that both TLR2 and TLR4 were involved in histone H4-mediated endothelial cell activation (**Supplementary Figure 5**) as has been previously reported (44). Inflammatory cytokines (IL-8 and MCP-1) also increased in HUVEC supernatants upon exposure to histone H4 and were subsequently suppressed by defibrotide (**Supplementary Figure 6**). Similar patterns were also observed for TF gene expression (**Figure 4D**) and enzymatic activity (**Supplementary Figure 7**). Given these findings, along with the RNA sequencing data, we investigated whether defibrotide might be working to counterbalance intracellular signaling pathways associated with TLRs or TNF signaling. However, in our hands, defibrotide showed only mild protection when HUVECs were activated by TNF-α and little to no protection when HUVECs were activated by lipopolysaccharide (**Supplementary Figure 8**). We therefore next tested whether defibrotide might work through direct engagement with histone H4 in the extracellular space. Indeed, using an electrophoretic mobility shift assay (EMSA) (45), we found evidence of a direct interaction between histone H4 and defibrotide in that histone H4 could retard the migration of defibrotide (a mixture of oligonucleotides) through an agarose gel (**Figure 4E**). In contrast, histone H4 had no impact on the migration of bovine serum albumin. A strong interaction between histone H4 and defibrotide was also confirmed by surface plasmon resonance. Assuming an average molecular weight of defibrotide as 16.5 kilodaltons, the equilibrium dissociation constant (K_D_) between defibrotide and histone H4 was calculated as 53.5 nM (**Figure 4F**).

**Figure 4:**
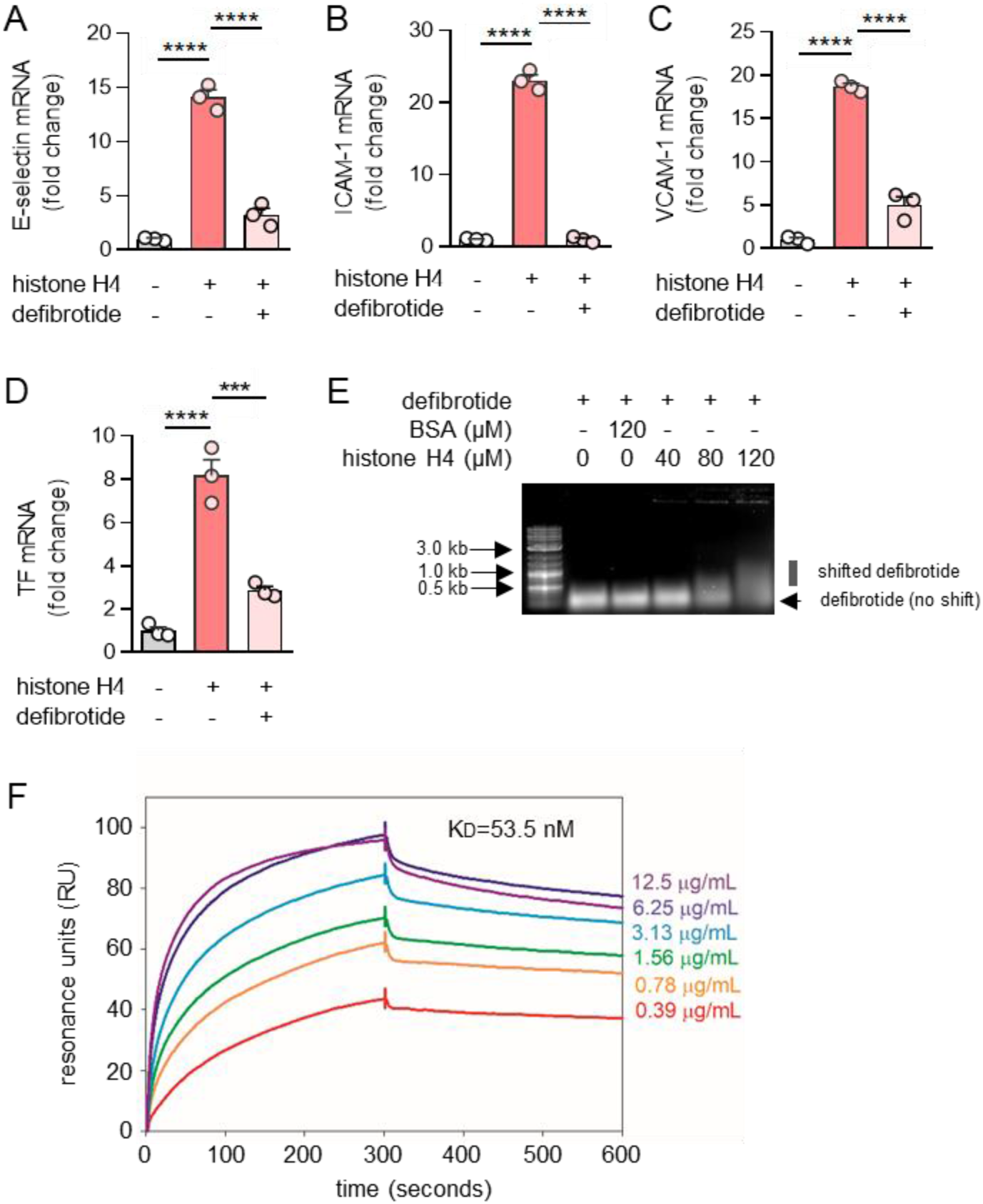
Defibrotide abolishes HUVEC activation by extracellular histone H4. **A-D,** HUVECs were pretreated with defibrotide (10 μg/ml) for 30 minutes, followed by recombinant histone H4 (25 μg/ml) for 4 hours. E-selectin (A), ICAM-1 (B), VCAM-1 (C), and tissue factor (D) mRNA levels were determined by qPCR. Mean ± standard deviation is presented for one representative experiment out of three independent experiments, all with similar results; “***p < 0.001, ****p < 0.0001 by one-way ANOVA corrected by Dunnett’s test. **E,** Defibrotide, and histone H4 were incubated at 37°C for 30 minutes and then resolved on a 0.5% agarose gel. **F,** Surface plasmon resonance assay characterizing the binding kinetics of defibrotide to histone H4. The profile of defibrotide at gradient concentrations (from 0.39 μg/ml to 12.5 μg/ml) flowing over histone H4 protein immobilized on a NiNTA chip are shown. The calculated dissociation constant (K_D_) is labeled.

### Defibrotide strongly protects endothelial cells against histone H4-induced cell death

The above assays were focused on relatively short cell culture times, typically 6 hours. However, we questioned whether the impact of defibrotide on histone H4-mediated HUVEC activation would persist over longer periods of time. As reported previously, histones go beyond endothelial cell activation and become cytotoxic upon prolonged exposure in culture (22, 46). Indeed, we found remarkable protection of cell viability by defibrotide over a 24-hour period (**Figure 5A**). We additionally found that 30-minute pretreatment with defibrotide was not absolutely necessary, as adding defibrotide one hour (but not later) after histone H4 preserved at least some of the protective effects (**Figure 5B**). A similar protective effect of defibrotide on cell viability was also observed when HMVECs were cultured together with histone H4 (**Supplementary Figure 9**). To further confirm these findings, we varied the experiment by introducing kinetic monitoring of surface phosphatidylserine exposure as measured by Annexin V binding. We found a dose-dependent relationship between histone H4 and Annexin V binding (**Supplementary Figure 10**) and found strong and stepwise protection when HUVECs were also cultured with defibrotide concentrations ranging from 10-40 μg/ml (**Figure 5C**). As previous work has revealed that defibrotide acts as an adenosine receptor agonist in some contexts (47), we asked whether adenosine A_2A_ or A_2B_ receptor antagonists could abolish the protective effects of defibrotide. We did not, however, find any role for the A_2A_ antagonist SCH 58261 or the A_2B_ antagonist PSB 603 in negating defibrotide’s protection against Annexin V binding (**Supplementary Figure 11**). We also assessed wortmannin, which has been reported to inhibit defibrotide uptake by endothelial cells (48), but again did not see an effect in our system **(Supplementary Figure 11)**. We did though find increased levels of both IL-1β and IL-18 in culture supernatants of HUVECs stimulated by histone H4, both of which were reduced by defibrotide (**Figure 6A-B**), supporting the idea that histone H4-mediated cell death may be on the spectrum of pyroptosis. To verify this hypothesis, gasdermin D (GSDMD, a critical protein mediating pyroptosis) and caspase 3 were characterized in HUVEC protein lysates cultured with either histone H4 or an apoptosis inducer staurosporine. In the histone H4-cultured HUVEC lysates, we found decreased expression of full-length GSDMD but increased expression of cleaved GSDMD, indicating that the type of HUVEC death triggered by histone H4 is pyroptosis, but not apoptosis (**Figure 6C**). To further substantiate these data, we also assessed translocation and subsequent release of the alarmin HMGB1, which is known to track with inflammatory forms of cell death including pyroptosis (49). By microscopy, we observed the translocation of HMGB1 from nucleus to cytoplasm upon exposure of HUVECs to histone H4, with reversal of this effect by defibrotide (**Figure 6D**). Measurement of HMGB1 in culture supernatants mirrored these findings, with histone H4 triggering HMGB1 release and defibrotide neutralizing that effect (**Figure 6E**). We also asked whether defibrotide can bind HMGB1, which is, like histone H4, a potentially cytotoxic cationic protein. Consistent with our hypothesis, an EMSA experiment suggested a direct interaction between HMGB1 and defibrotide (**Supplementary Figure 12**), hinting at another mechanism by which defibrotide might restrain inflammation downstream of histones and NETs. Taken together, these data demonstrate that longer-term exposure of HUVECs to histone H4 triggers pyroptosis and that defibrotide’s anti-histone effects are sustained in culture for up to 24 hours.

**Figure 5:**
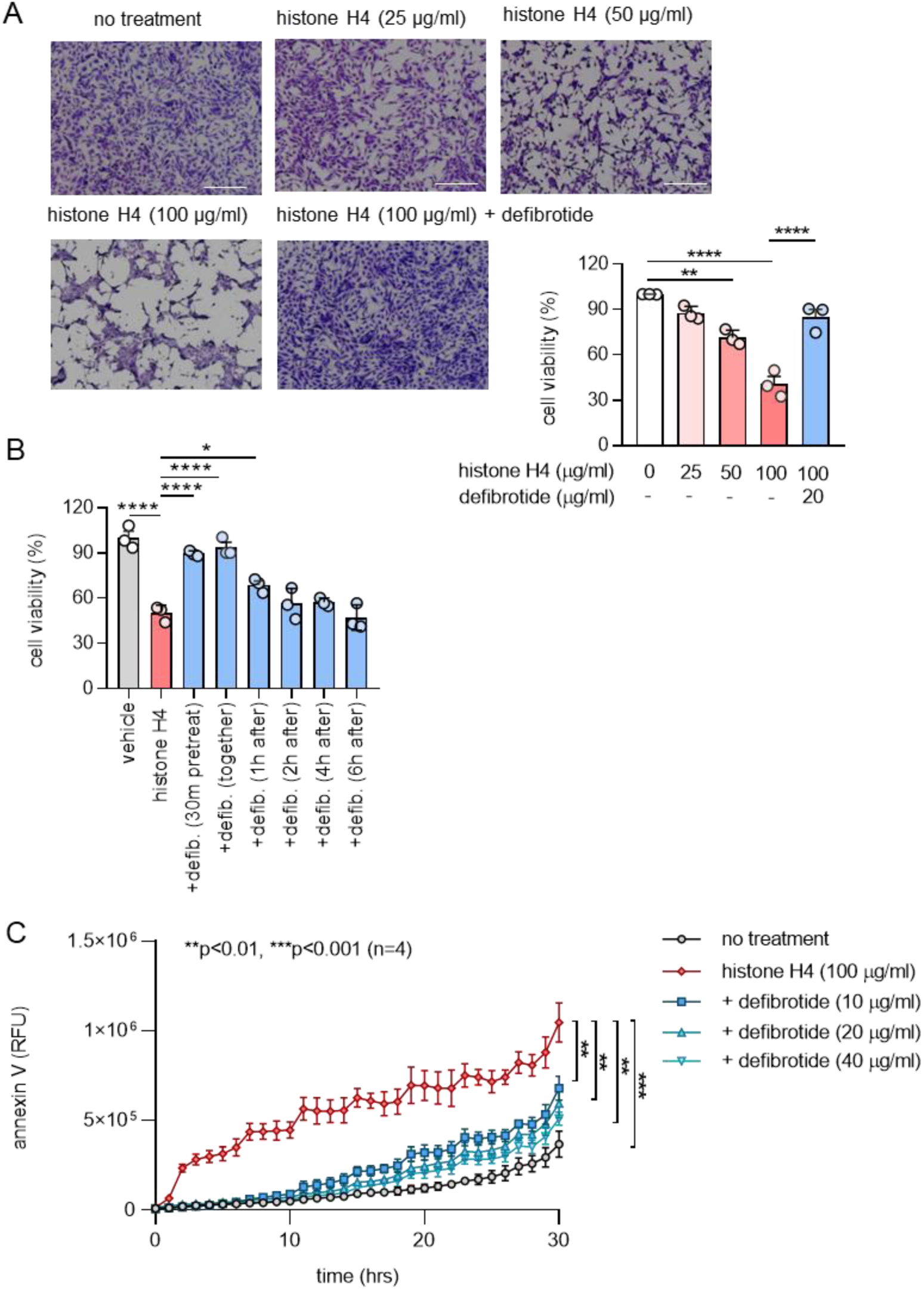
Defibrotide protects HUVECs from histone H4-mediated cell death. **A,** HUVECs were treated with different doses of histone H4 (0, 25, 50, and 100 μg/ml) in the presence or absence of defibrotide (20 μg/ml). After 24 hours, HUVECs were stained with crystal violet solution for 10 minutes, and absorbance was measured at 570 nm to determine cell viability. Mean ± standard deviation for three independent experiments, along with representative images, are presented; ***p<0.001 and ****p<0.0001 by one-way ANOVA corrected by Tukey’s multiple comparisons test. Scale bars are 500 microns. **B**, HUVECs were treated with histone H4 (25 μg/ml) in the presence or absence of defibrotide (20 μg/ml, added at different time points relative to histone H4). After 24 hours, HUVECs were stained with crystal violet solution for 10 minutes, and absorbance was measured at 570 nm to determine cell viability. Mean ± standard deviation for three independent experiments is presented; *p<0.05, ***p<0.001 and ****p<0.0001 by one-way ANOVA corrected by Turky’s test**. C,** HUVECs were treated with histone H4 and different doses of defibrotide in the presence of Annexin V red agent. The plate was imaged every hour using the IncuCyte^®^ S3 timelapse microscope for 30 hours. Mean ± standard deviation for three independent experiments is presented; **p<0.01 and ***p<0.001 by two-way ANOVA corrected by Dunnett’s test.

**Figure 6:**
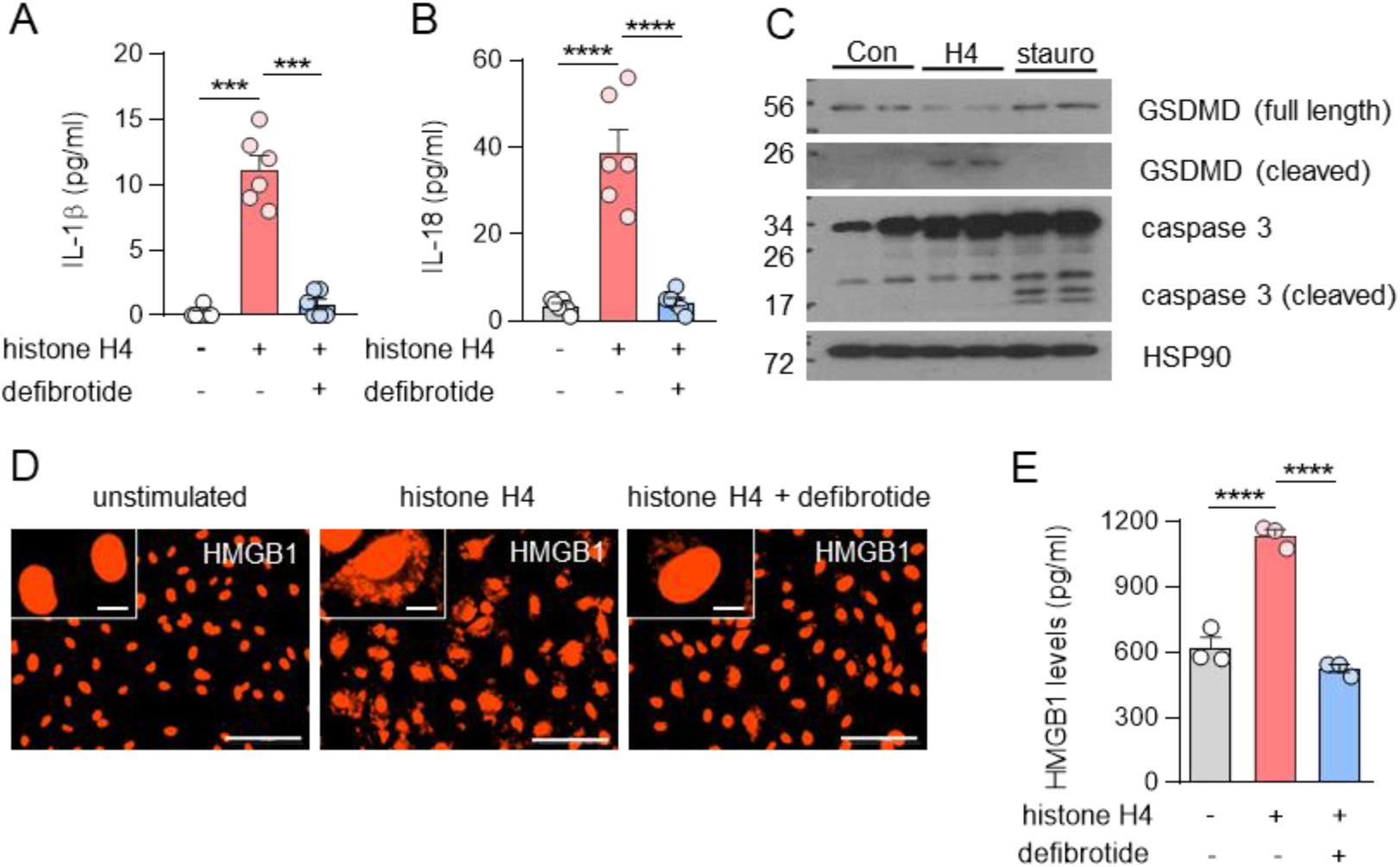
Defibrotide protects HUVECs from histone H4-mediated pyroptosis. **A-B,** HUVECs were treated with histone H4 (100 µg/ml) ± defibrotide (20 µg/ml) for 4 hours. The concentrations of IL-1β (C) and IL-18 (D) were determined in supernatants (n=6 independent experiments); ***p<0.001 and ****p<0.0001 by one-way ANOVA corrected by Dunnett’s test. Data was presented as mean ± standard deviation. **C,** Immunoblotting detection of activated gasdermin D (GSDMD) and caspase 3 in cell lysates. HUVECs were treated with histone H4 (100 µg/ml) or staurosporine (50 nM) for 6 hours before collecting the cell lysates. Con=control, H4=histone H4, stauro=staurosporine. **D-E,** HUVECs were treated as in panels A-B, and HMGB1 translocation (D) and secretion (E) were determined by microscopy and supernatant ELISA, respectively (n=3 independent experiments); ****p<0.0001 by one-way ANOVA corrected by Dunnett’s test. Scale bars are 100 microns (primary image) and 10 microns (inset). Data was presented as mean ± standard deviation.

### Defibrotide counters histone-accelerated venous thrombosis in mice

To determine the potential *in vivo* relevance of these findings, we employed a model of venous thrombosis in which a constricting ligature is fixed around the inferior vena cava (IVC) and then the presence and potential characteristics of thrombosis are assessed 24 hours later (**Figure 7A**) (6, 50). First, we asked whether thrombus accretion was impacted by injection of calf thymus histones, and indeed found this to be the case (**Figure 7B-C**); at the same time, extensive review of kidney sections did not reveal spontaneous structures in glomeruli or vessels suggestive of thrombi (demonstrating that a second hit was needed in addition to histone injection). We also did not find that histone injection boosted the levels of myeloperoxidase-DNA complexes (NET remnants) in blood (data not shown). As part of these experiments, we also administered defibrotide intravenously shortly after injection of the histones. With this approach, both thrombus accretion (**Figure 7B-C**) and thrombus length (**Supplementary Figure 13**) were reduced essentially to the levels seen in control mice. In support of endothelial cell activation contributing to the histone-accentuated thrombosis phenotype, both soluble E-selectin and soluble P-selectin tracked closely with thrombus accretion (**Figure 7D-E**), as did infiltration of leukocytes, whether scored as Ly6G+ (neutrophils) or CD45+ (most leukocytes) (**Figure 7F-H**). In support of this concept, there was a strong correlation between either soluble E-selectin or soluble P-selectin and thrombus size (**Supplementary Figure 14**). Taken together, these data confirm the proinflammatory and prothrombotic impact of histones *in vivo* and demonstrate that defibrotide has the potential to neutralize these properties.

**Figure 7:**
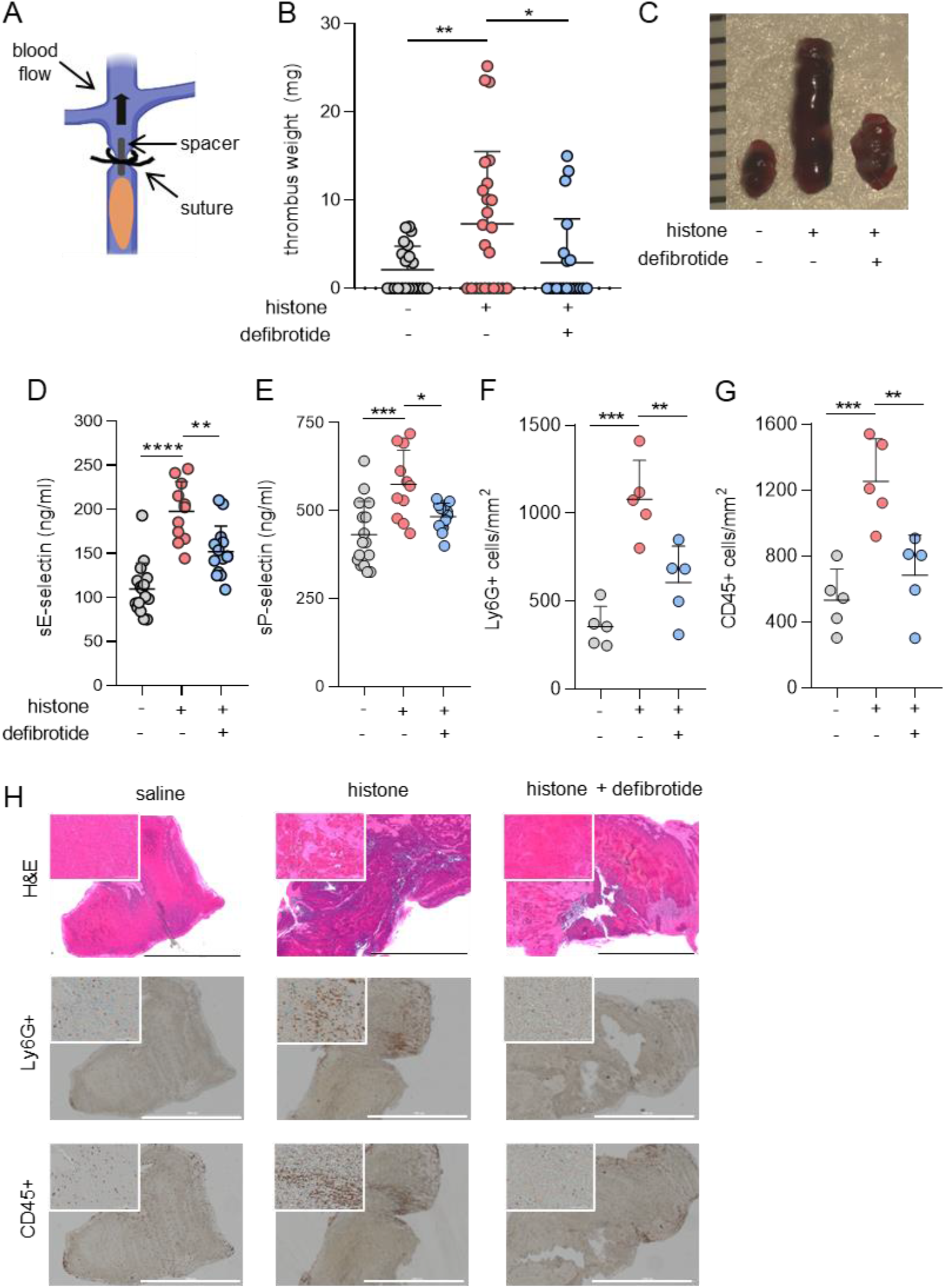
Defibrotide alleviates histone-mediated endothelial activation and venous thrombosis in mice. **A,** Thrombus initiation in the IVC via placement of a fixed suture over a spacer that was subsequently removed. **B,** Mice were injected with either histone (10 mg/kg) or saline via tail vein 1 hour prior to surgery. Meanwhile, defibrotide (150 mg/kg) or saline was administered by retro-orbital injection 24 hours prior to surgery and then immediately following closure of the abdomen. Thrombus weight was determined 24 hours later. Scatter plots are presented with each data point representing a unique mouse (horizontal bars=means; *p<0.05 and **p<0.01 by Kruskal-Wallis test followed by Dunn’s multiple comparison test. Data was presented as mean ± standard deviation **C,** Representative thrombi from the experiments presented in panel B with rulers measuring thrombi in millimeters. **D-E,** Serum samples from the experiments presented in panel B were tested for soluble E-selectin (D) and soluble P-selectin (E) by ELISA; *p<0.05, **p<0.01, and ***p<0.001 by one-way ANOVA corrected by Dunn’s multiple comparison test. Data was presented as mean ± standard deviation **F-H,** Thrombus sections from panel B were stained for Ly6G and CD45-positive cells. Positively-stained cells were quantified in four randomly selected fields for each thrombus. **p<0.01 and ***p<0.001 by one-way ANOVA corrected by Dunn’s multiple comparison test. Scale bars=1000 microns. Data is presented as mean ± standard deviation.

## DISCUSSION

As evidence continues to implicate NETs and NET-derived histones in the pathophysiology of disease states ranging from infection (including COVID-19) to autoimmunity to cancer (51), the search for NET-targeting therapeutics takes on additional importance. Here, we explored the extent to which an FDA-approved drug defibrotide might protect endothelial cells from NETs and extracellular histones. We found defibrotide to counteract endothelial cell activation and hypercoagulability triggered by NETs, and histone H4. Mechanistically, our evidence points to a direct interaction between defibrotide and cationic proteins such as histone H4 as an important aspect of these protective effects.

Polyanionic substances naturally exist in the extracellular environment where they play a variety of biological roles (52). Unfractionated heparin and suramin are examples of how polyanionic drugs may be leveraged clinically with the potential to bind cationic microbe-derived proteins as well as cationic tumor cytokines/receptors for treatment of infectious diseases and cancer, respectively (53–57). In previous work, defibrotide’s polyanionic properties have been shown to include binding with high affinity to specific heparin-binding proteins including basic fibroblast growth factor (bFGF) (40). Interestingly, defibrotide (oligonucleotides) and heparins (proteoglycans) share similarities in charge distributions and binding patterns (58).

Histones bind DNA tightly mainly due to charge-charge interactions, with a possible role for specific DNA sequence motifs (59). Our results found a strong interaction between histone H4 and defibrotide, which was very resistant to dissociation. Considering defibrotide is a natural product (i.e., not produced by a DNA synthesizer), the possibility of any specific sequence dominating its effect is low. We therefore speculate that the main binding force between histone H4 and defibrotide also comes from charge-charge interactions. An interesting unknown is the extent to which the degradation of defibrotide *in vivo* may be delayed upon binding histone H4. Typically, phosphodiester oligonucleotides would be rapidly degraded in plasma; however, we found a protective role in an animal model over 24 hours without the need for redosing. A deeper understanding of these *in vivo* properties should be a priority for future research.

As a major component of NETs, histones are one factor that contributes to vascular dysfunction during sepsis where they trigger neutrophil migration, endothelial injury, hemorrhage, and thrombosis (17). Compared with other histones, histone H4 has the strongest impact on platelets, enhancing thrombin generation and accelerating thrombosis (60). Histone H4 has also been reported as the major histone mediator of membrane lysis of smooth muscle cells and arterial tissue damage and inflammation in atherosclerosis (61). Our study has now also revealed that neutralizing histone H4 significantly mitigates NET-mediated activation of HUVECs. Whether defibrotide preferentially neutralizes histone H4 as compared with other histones is an area deserving of further research.

Multiple organ dysfunction syndrome (MODS) is widely considered to be the leading cause of morbidity and mortality for patients admitted to an intensive care unit, where it encompasses heterogeneous disease states such as sepsis, shock, trauma, severe burn, and pancreatitis (62–65). Systemic inflammation and vascular coagulopathy account for the main pathological processes of MODS (66), and of course also characterize aspects of COVID-19 (67–70) and the closely related catastrophic antiphospholipid syndrome (71). In MODS, endothelial cell activation is considered a precursor to tissue damage and end-organ dysfunction with upregulation of adhesion molecules triggered by cytokines, microbial proteins, and various cationic proteins from necrotic cells (72). One recent study evaluated circulating histones in a cohort of 420 ICU patients with sepsis, severe trauma, or severe pancreatitis and identified circulating histones as major mediators of MODS in these patients (73). An important future direction of this work will be to characterize the role of histones and defibrotide in the context of *in vivo* models that interrogate the microvasculature, where much of the pathology of MODS resides. One may then be able to consider whether administration of defibrotide in an early phase of MODS might neutralize cationic proteins such as histones to stabilize the endothelium and break the vicious thromboinflammatory cycle. Indeed, a number of clinical trials focused on defibrotide therapy for COVID-19 are currently underway or recently completed (NCT04530604, NCT04335201, NCT04348383, NCT04652115). These and potentially other future trials should help elucidate the extent to which defibrotide and other histone-neutralizing agents may have a role in combatting NET-mediated disease states.

## METHODS

### Cell culture and reagents

Human umbilical vein endothelial cells (HUVECs) and human dermal microvascular endothelial cells (HMVECs) purchased from ATCC were cultured in EBM supplemented with EGM-2MV singleQuots (Lonza) without hydrocortisone in 0.2% gelatin-coated tissue culture plates. All experiments were performed using HUVECs of passage 6 or lower. Recombinant histone H4 was purchased from Cayman (10264) for *in vitro* experiments. Histone from calf thymus was purchased from Sigma (10223565001). Anti-histone H4 was from Cell Signaling Technology (2592). TLR2 inhibitor C29 was from Medchemexpress (HY-100461) and TLR4 inhibitor TAK 242 was from Sigma (614316). Citrullinated histone H4 (17927), SCH 58261 (19676), PSB 603 (25637), and wortmannin (10010591) were purchased from Cayman.

### NET isolation

Neutrophils were isolated from healthy volunteers. Neutrophil extracellular traps (NETs) were stimulated with 500 nM phorbol 12-myristate 13-acetate (PMA) or 10 µM calcium ionophore A23187 and purified as described previously (74).

### Quantitative polymerase chain reaction (qPCR)

Total RNA was isolated using Direct-zol RNA MiniPrep kit (Zymo Research) according to manufacturer’s instructions. 200 ng of RNA from each sample was reverse-transcribed using random hexamer primed single-strand cDNA (10 min at 25 °C, 15 min at 42 °C, 5 min at 99 °C) by MMLV Reverse Transcriptase (Life technologies). cDNA was amplified using Fast SYBR Green Mastermix (Life Technologies) on a ViiA7-Realtime qPCR System (Life Technologies). Expression level of mRNAs were normalized to β-actin. All gene primers were purchased from Qiagen.

### Neutrophil adhesion assay

Monolayer HUVECs were cultured with or without NETs for 4 hours. Isolated fresh neutrophils were labeled with calcein-AM (C1430, Thermo) for 30 minutes at 37°C, and then 6×10^5^ neutrophils per well were added to the washed (RPMI + 3% BSA) monolayer for 20 minutes. After washing with pre-warmed HBSS, adherent neutrophil fluorescence was measured with a Cytation 5 Cell Imaging Multi-Mode Reader (BioTek) at 485 and 535 nm (excitation and emission wavelengths, respectively).

### TF activity

Cell lysates were prepared with 150 µl 15 mM octyl-ß-D-glycopyranoside (Sigma) for 15 min at 37°C. TF activity was measured using TF Human Chromogenic Activity Assay Kit (ab108906, Abcam) according to the manufacturer’s instructions.

### Permeability assay

Permeability was assessed by measuring the passage of horseradish peroxidase (HRP) through endothelial cell monolayers in a Transwell system (Cell Biologics). Briefly, HUVECs were plated at 50,000 cells/ml in the Transwells and allowed to grow to confluence, and then cultured in EBM-2 media with 1% fetal bovine serum (FBS) in the upper and lower chambers. Treatments, including NETs (1 μg DNA content/ml) and/or defibrotide (10 μg/ml), were added to the upper chambers along with HRP, and aliquots of the media in the lower chambers were collected at various time points. The amount of HRP was quantified by the addition of 3,3’,5,5’-tetramethylbenzidine and stop solution. Absorbance was read at 450 nm in a plate reader.

### In-cell ELISA

Confluent monolayers of HUVECs in 96-well microplates were incubated with NETs for 6 hours. Some cultures were additionally supplemented with defibrotide. Cells were fixed by adding an equal volume of 8% paraformaldehyde for 30 minutes. Blocking was with 2x blocking solution (ab111541, Abcam) at room temperature for 2 hours. After washing with PBS, cells were incubated with 5 µg/ml primary mouse anti-human antibodies against E-selectin (BBA26, R&D), VCAM-1 (BBA5, R&D), or ICAM-1 (ab2213, Abcam) at 4°C overnight. Next, 100 µl of diluted horseradish peroxidase conjugated rabbit anti-mouse IgG (1:2000, Jackson ImmunoResearch) in 1x blocking solution was added and incubated at room temperature for 1 hour. After washing thoroughly with PBS, 100 µl of TMB substrate was added, and blue color development was measured at OD 650 nm with a Cytation 5 Cell Imaging Multi-Mode Reader (BioTek). The signals were corrected by subtracting the mean signal of wells incubated in the absence of the primary antibody.

### RNA-sequencing

Total RNA from cells was isolated using RNeasy Plus Mini Kit (74134, Qiagen) according to manufacturer’s instructions. Sequencing was performed by the UM Advanced Genomics Core, with libraries constructed and subsequently subjected to 150 paired-end cycles on the NovaSeq-6000 platform (Illumina). FastQC (v0.11.8) was used to ensure the quality of data, and adapter sequences were trimmed from raw reads using Cutadapt (v2.3) prior to alignment. Reads were mapped to the reference genome GRCh38 (ENSEMBL) using STAR (v2.6.1b) and assigned count estimates to genes with RSEM (v1.3.1). Alignment options followed ENCODE standards for RNA-seq. FastQC was used in an additional post-alignment step to ensure that only high-quality data were used for expression quantitation and differential expression. Differential expression data were pre-filtered to remove genes with 0 counts in all samples. Differential gene expression analysis was performed using DESeq2, using a negative binomial generalized linear model (thresholds: linear fold change >1.5 or <-1.5, Benjamini-Hochberg FDR (Padj) <0.05). Plots were generated using variations of DESeq2 plotting functions and other packages with R version 3.3.3. Functional analysis, including candidate pathways activated or inhibited in comparison(s) and GO-term enrichments, was performed using iPathway Guide (Advaita). RNA sequencing data discussed in this publication have been deposited in NCBI’s Gene Expression Omnibus and are accessible through GEO Series accession number GSE179828 (https://www.ncbi.nlm.nih.gov/geo/query/acc.cgi?acc=GSE179828).

### Quantification of cytokines

Cytokines were quantified in supernatants using human IL-1β DuoSet ELISA kit (DY201, R&D systems), human IL-18 ELISA Kit (7620, MBL International), human IL-8 DuoSet ELISA kit (DY208, R&D systems), and human MCP-1 DuoSet ELISA kit (DY279, R&D systems), according to the manufacturers’ instructions.

### Electrophoretic mobility shift assay (EMSA)

10 µg of defibrotide was incubated with various concentrations (40 µM, 80 µM, 120 µM) of histone H4 in serum-free RPMI for 1 hour at 37°C to form complexes. 120 µM bovine serum albumin was used as a negative protein control. Complexes were then run on a 0.5% agarose gel stained with SYBR safe (Invitrogen) for 30 minutes. The gel was imaged on a Typhoon FLA 7000 biomolecular imager (GE Healthcare).

### Surface Plasmon Resonance (SPR) assay

SPR studies were performed using the Biacore T200 with His-tagged histone H4 coupled to a NiNTA chip. Defibrotide in a series of concentrations from 0.39 µg/ml to 12.5 µg/ml was injected over the sensor chip at room temperature, using a running biffer of 50 mM Hepes Naoh pH 7.4, 100 mM NaCl, and 0.002% surfactant P-20. Resonance was corrected for background using a reference cell without histone H4, and curves were blank subtracted using data acquired with running buffer only. Data were analyzed using BIA Evaluation software (GE Healthcare Bio-Sciences, Pittsburgh, PA) to determine binding affinity at steady state. Data shown are representative of 3 independent experiments.

### Crystal violet viability staining

Cell viability was tested by crystal violet staining as reported previously(75).

### Annexin V staining

Cells were seeded into a 96-well plate and allowed to adhere overnight. Annexin V reagent (Incucyte Annexin V Green Reagent for Apoptosis; Essen Bioscience, final dilution of 1:200) was added together with histone H4 with or without defibrotide on the following day. Annexin V staining was monitored with the IncuCyte^®^ S3 microscopy system every 1 hour for 30 hours. Excitation and emission wavelengths were 490 nm and 515 nm, respectively. Images were collected by a Nikon 20x objective. IncuCyte^®^ S3 integrated software (Essen Bioscience) was used to minimize background fluorescence and quantify fluorescent objects.

### Immunoblotting analysis

Cells were harvested and homogenized with lysis buffer containing 2% SDS, 50 mM Tris-HCl (pH 6.8), 10 mM DTT, 10% glycerol, 0.002% bromphenol blue, and freshly added protease inhibitors. Immunoblotting experiments were performed using specific antibodies. Antibodies used in this study were against caspase 3 (catalog 9662, Cell Signaling Technology), full-length gasdermin D (catalog 96458, Cell Signaling Technology), cleaved N-terminal gasdermin D (catalog ab215203, Abcam) and Hsp90 (catalog sc-7947, Santa Cruz).

### Detection of HMGB1

For immunofluorescence microscopy, 1×10^5^/well HUVECs were seeded onto coverslips coated with 0.2% gelatin the day before experiments. The HUVECs were treated with 100 μg/ml histone H4 in the presence or absence of defibrotide for 24 hours. Cells were fixed with 1% paraformaldehyde for 10 minutes, permeabilized with 0.5% TritonX-100 for 10 minutes, and blocked with 5% FBS for 30 minutes. Then the cells were intracellularly stained with 5 µg/ml anti-HMGB1 Alexa Fluor® 594 (clone 3E8, Biolegend) in blocking buffer overnight at 4°C. Images were collected with a Cytation 5 Cell Imaging Multi-Mode Reader (BioTek). HMGB1 was quantified in supernatants using the HMGB1 ELISA Kit (NBP2-62766, Novus) according to the manufacturer’s instructions.

### Mouse models of venous thrombosis

Male C57BL/6 mice were purchased from The Jackson Laboratory (000664), and used at approximately 10 weeks of age. Large-vein thrombosis was modeled as we have described previously(6). Mice were injected with either histone (10 mg/kg) or saline 1 hour prior to surgery via tail vein. Defibrotide (150 mg/kg) or an equal volume of saline were administered intravenously via retro-orbital injection. The first dosage of defibrotide was given 24 hours prior to surgery, and the second dose was delivered just after closure of the abdomen. Thrombus was determined 24 hours later.

### Quantification of mouse soluble E-selectin and P-selectin

Soluble E-selectin and P-selectin were quantified in mice sera using the mouse E-selectin Duoset ELISA (DY575, R&D system) and mouse P-selectin Duoset ELISA (DY737, R&D system) according to the manufacturer’s instructions.

### Statistical analysis

Data analysis was performed with GraphPad Prism software version 8. For continuous variables, group means were compared by either t-test (two groups) or one-way ANOVA (more than two groups); correction for multiple comparisons was by Dunnett’s, Sidak’s, or Tukey’s method. For two independent variables, group means were compared by two-way ANOVA (more than two groups); correction for multiple comparisons was by Dunnett’s method. Correlations were tested by Pearson’s correlation coefficient. Statistical significance was defined as p<0.05.

### Study approval

Neutrophils were isolated from healthy volunteers recruited through an IRB-approved advertisement (HUM00044257). All mouse experiments were approved by the University of Michigan IACUC.

## Data Availability

Upon publication, RNA sequencing data will be uploaded to a public server. Other data will be made available by email to the corresponding author.

## ACKNOWLEDGEMENTS

The authors thank Doruk Erkan (Hospital for Special Surgery) for helpful discussions regarding potential roles of defibrotide in thromboinflammatory diseases, and thank Kelsey Rampalski for technical assistance with sectioning and histology. YZ was supported by a career development grant from the Rheumatology Research Foundation. YK was supported by the Intramural Research Program of the NIH and NHLBI, Lasker Foundation, Falk Medical Research Trust Catalyst Award, and the JOBST-American Venous Forum Award. JHM and SAS were supported by the NIH (R35 HL135823). JSK was additionally supported by grants from the NIH (R01HL115138), Burroughs Wellcome Fund, Rheumatology Research Foundation, and Lupus Research Alliance.

## AUTHORSHIP

HS, AAG, SS, QWY, DC, SY, RAA, CL, GS, and P-ST conducted experiments and analyzed data. HS, YZ, YK, EAF, JDL, JHM, and JSK conceived the study and analyzed data. All authors participated in writing the manuscript and gave approval before submission.

## Supplementary Materials

**Supplementary Figure 1:**
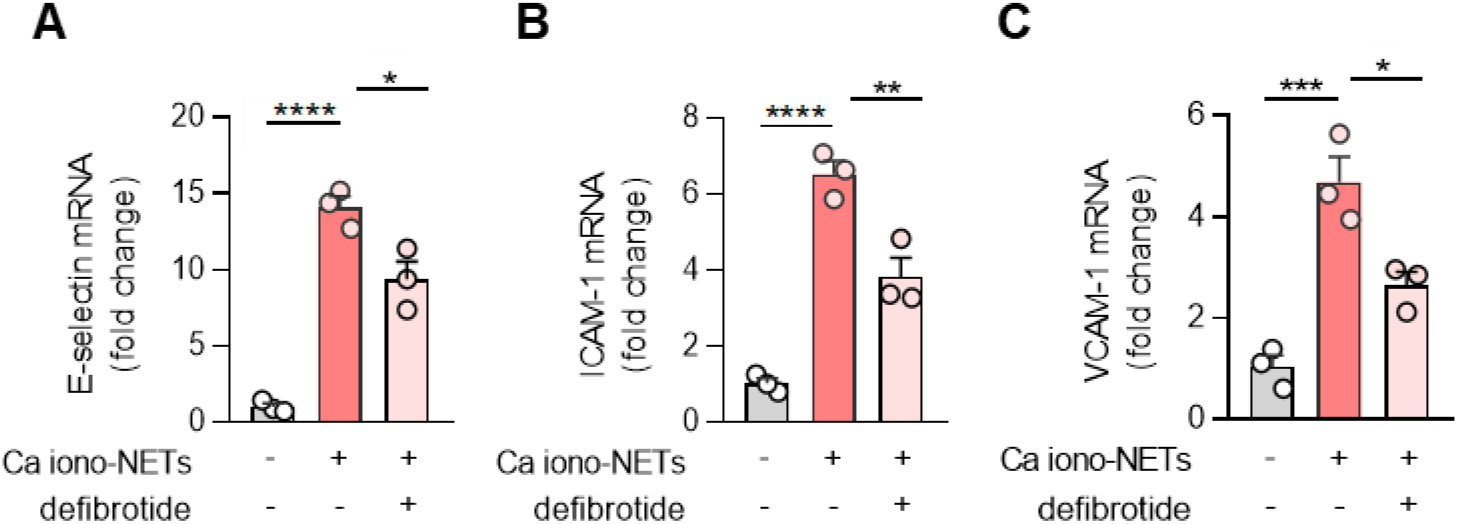
Defibrotide inhibits the activation of cultured HUVECs by calcium ionophore-induced NETs. **A-C,** HUVECs were pretreated with defibrotide (10 μg/ml) for 30 minutes, followed by isolated NETs (1 μg DNA content/ml triggered by 10 μM calcium ionophore) for 4 hours. E-selectin (A), ICAM-1 (B), and VCAM-1 (C) mRNA levels were determined by qPCR. Mean ± standard deviation is presented for one representative experiment out of three independent experiments, all with similar results; ****p<0.0001 by one-way ANOVA corrected by Dunnett’s test.

**Supplementary Figure 2:**
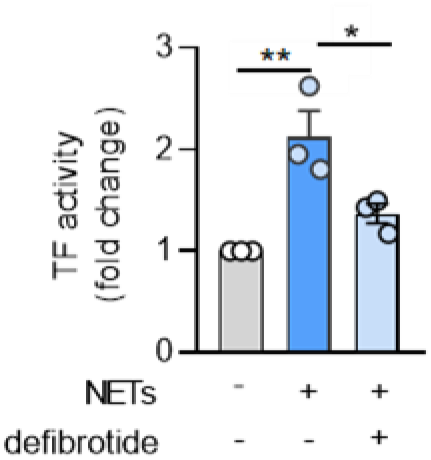
Defibrotide inhibits the tissue factor activity of cultured HUVECs by NETs. HUVEC lysates were prepared and tissue factor activity was determined as described in Methods. Mean ± standard deviations are presented for n=3 independent experiments; *p<0.05 and **p<0.01 by one-way ANOVA corrected by Dunnett’s test.

**Supplementary Figure 3:**
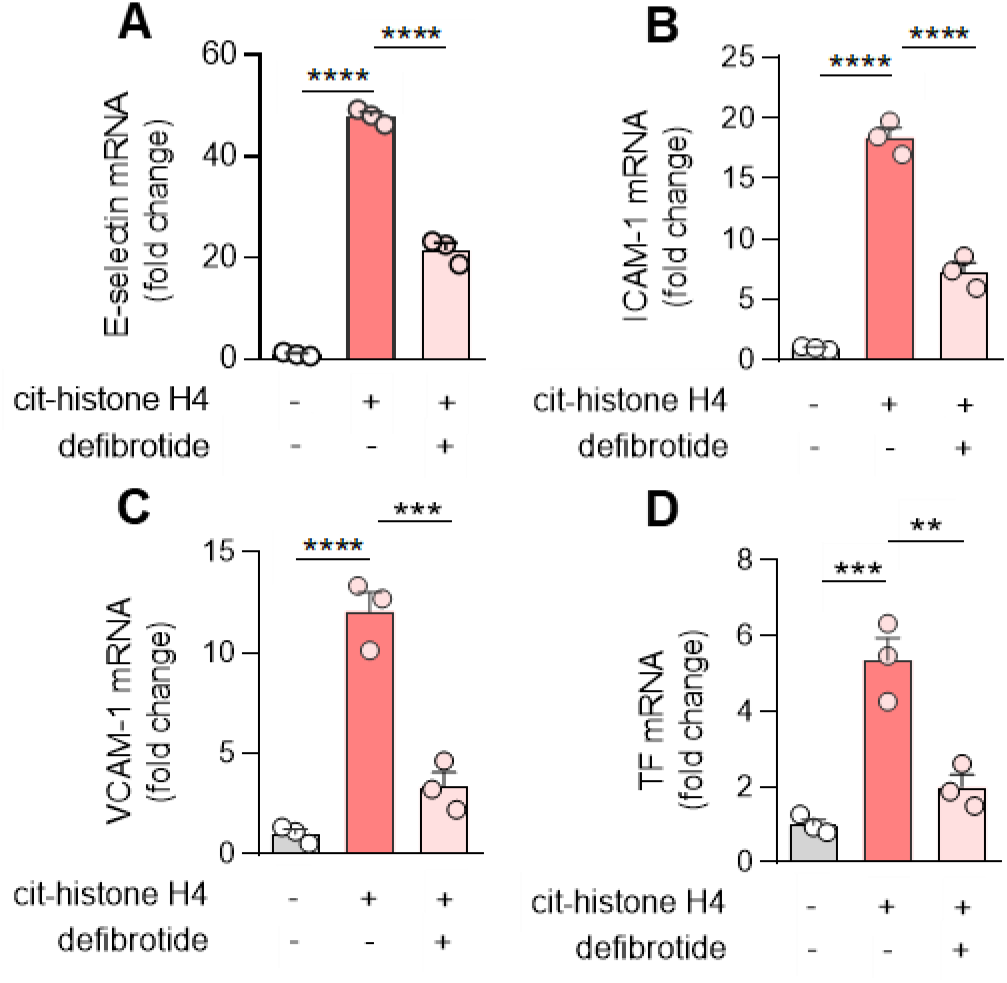
Defibrotide mitigates HUVEC activation by citrullinated histone H4 (cit-histone H4). **A-D,** HUVECs were pretreated with defibrotide (10 μg/ml) for 30 minutes, followed by recombinant citrullinated histone H4 (25 μg/ml) for 4 hours. E-selectin (A), ICAM-1 (B), VCAM-1 (C), and tissue factor (D) mRNA levels were determined by qPCR. Mean ± standard deviation is presented for one representative experiment out of three independent experiments, all with similar results; **p < 0.01, ***p < 0.001, ****p < 0.0001 by one-way ANOVA corrected by Dunnett’s test.

**Supplementary Figure 4:**
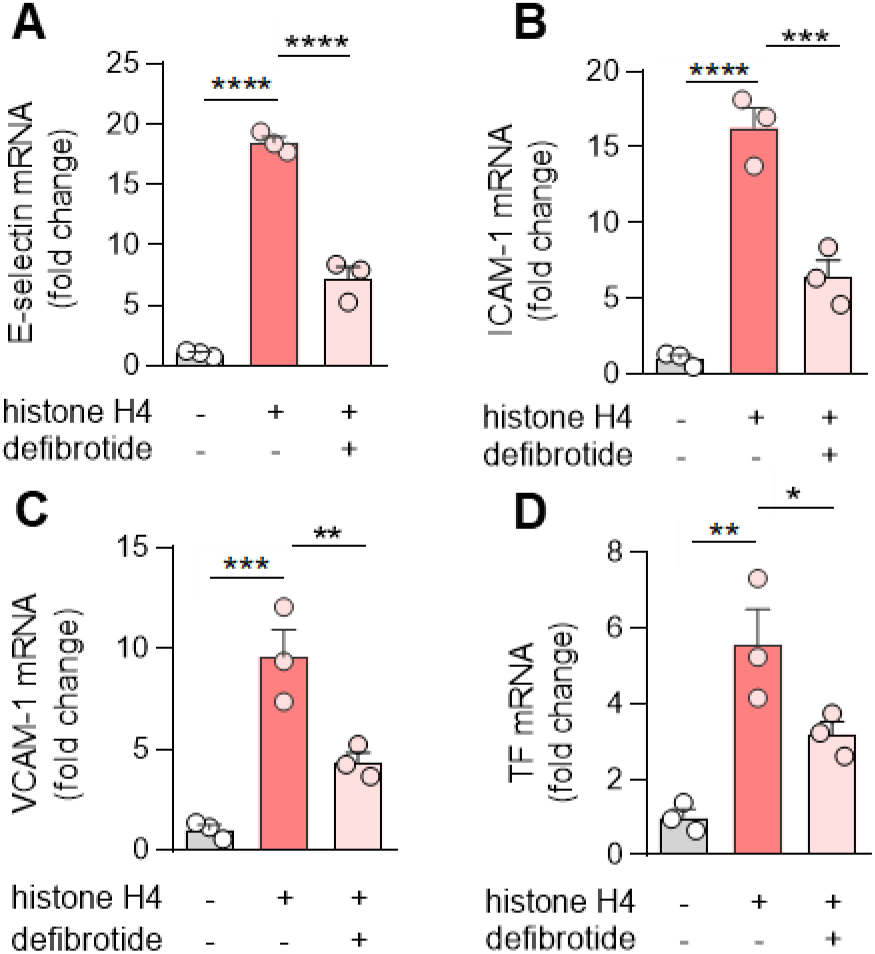
Defibrotide attenuates human dermal microvascular endothelial cell (HMVEC) activation by extracellular histone H4. **A-D,** HMVECs were pretreated with defibrotide (10 μg/ml) for 30 minutes, followed by recombinant histone H4 (25 μg/ml) for 4 hours. E-selectin (A), ICAM-1 (B), VCAM-1 (C), and tissue factor (D) mRNA levels were determined by qPCR. Mean ± standard deviation is presented for one representative experiment out of three independent experiments, all with similar results; *p < 0.05, **p < 0.01, ***p < 0.001, ****p < 0.0001 by one-way ANOVA corrected by Dunnett’s test.

**Supplementary Figure 5:**
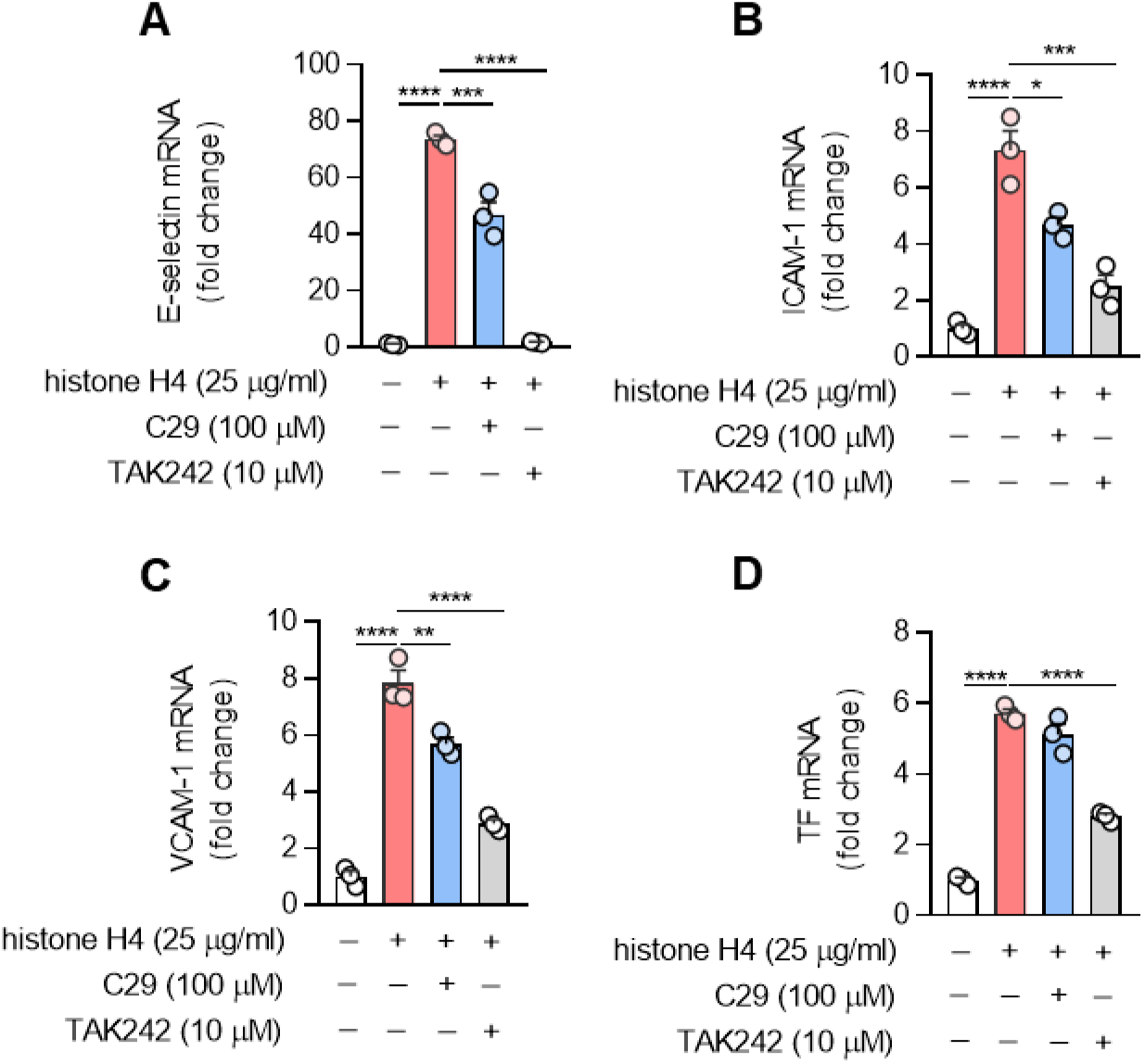
Toll-like receptor inhibitors mitigate the activation of cultured HUVECs by histone H4. **A-D,** HUVECs were pretreated with C29 (TLR2 inhibitor) or TAK242 (TLR4 inhibitor) for 1 hour, followed by the addition of histone H4 (25 μg/ml) for 4 hours. E-selectin (A), ICAM-1 (B), VCAM-1 (C) and tissue factor (D) mRNA levels were determined by quantitative PCR. Mean ± standard deviation is presented for one representative experiment out of three independent experiments, all with similar results; *p<0.05, **p<0.01, ***p<0.001, and ****p<0.0001 by one-way ANOVA corrected by Dunnett’s test.

**Supplementary Figure 6:**
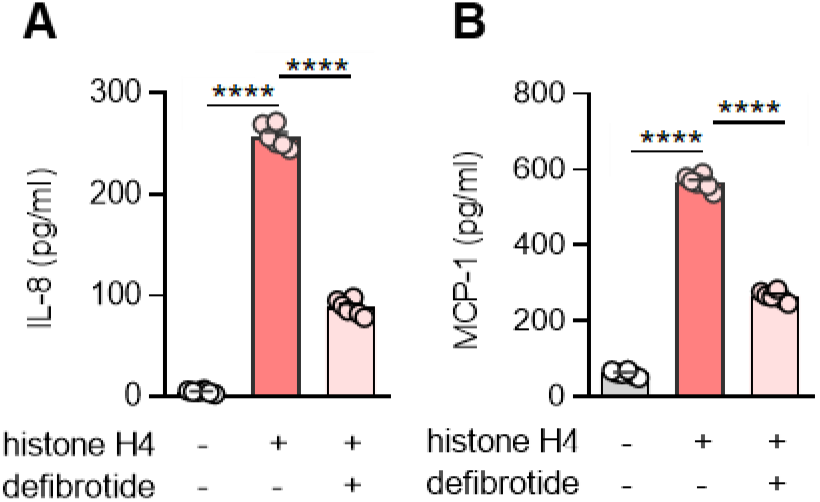
Defibrotide protects HUVECs from histone H4-mediated chemokine secretion. HUVECs were treated with histone H4 (25 µg/ml) ± defibrotide (10 µg/ml) for 4 hours. The concentrations of IL-8 (A) and MCP-1 (B) were determined in supernatants (n=6 independent experiments); ****p<0.0001 by one-way ANOVA corrected by Dunnett’s test.

**Supplementary Figure 7:**
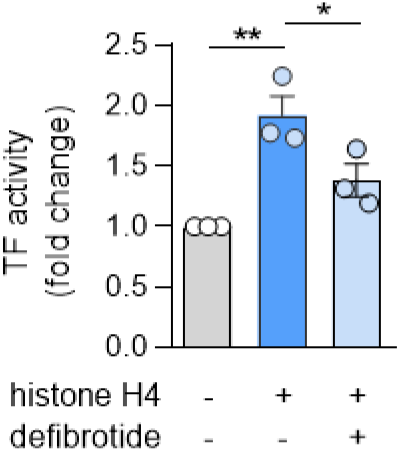
Defibrotide inhibits the tissue factor activity of cultured HUVECs by histone H4. HUVEC lysates were prepared and tissue factor activity was determined as described in Methods. Mean ± standard deviations are presented for n=3 independent experiments; *p<0.05 and **p<0.01 by one-way ANOVA corrected by Dunnett’s test.

**Supplementary Figure 8:**
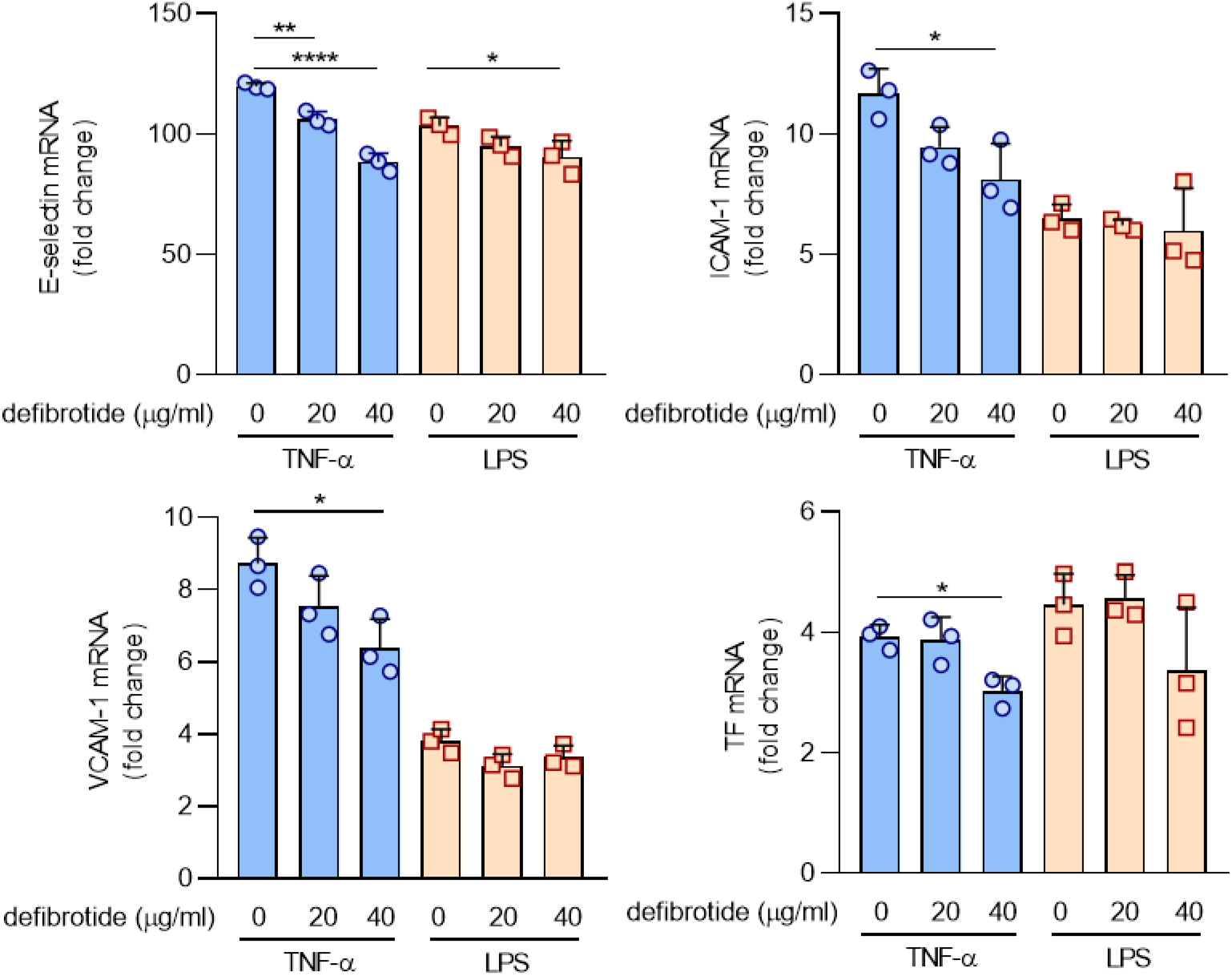
Defibrotide shows mild protection when HUVECs are activated by TNF-α and little to no protection when HUVECs are activated by lipopolysaccharide (LPS). **A-D,** HUVECs were pretreated with defibrotide (20 or 40 μg/ml) for 30 minutes, followed by TNF-α (20 nM) or LPS (1 µg/ml) for 4 hours. E-selectin (A), ICAM-1 (B), VCAM-1 (C), and tissue factor (D) mRNA levels were determined by qPCR. Mean ± standard deviation is presented for one representative experiment out of three independent experiments, all with similar results. *p<0.05, **p < 0.01, ****p < 0.0001 by one-way ANOVA corrected by Dunnett’s test.

**Supplementary Figure 9:**
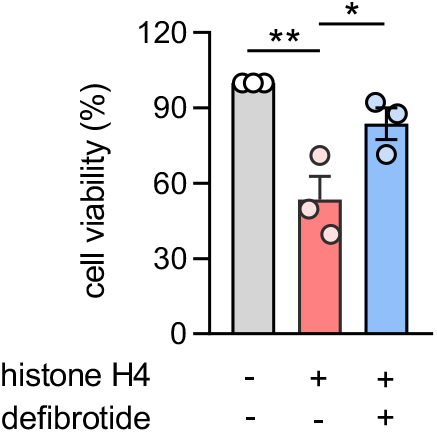
Defibrotide protects HMVECs from histone H4-mediated cell death. HMVECs were treated with histone H4 (25 μg/ml) in the presence or absence of defibrotide (20 μg/ml). After 24 hours, HVECs were stained with crystal violet solution for 10 minutes, and absorbance was measured at 570 nm to determine cell viability. Mean ± standard deviation for three independent experiments is presented; *p<0.05 and **p<0.01 by one-way ANOVA corrected by Dunnett’s test.

**Supplementary Figure 10:**
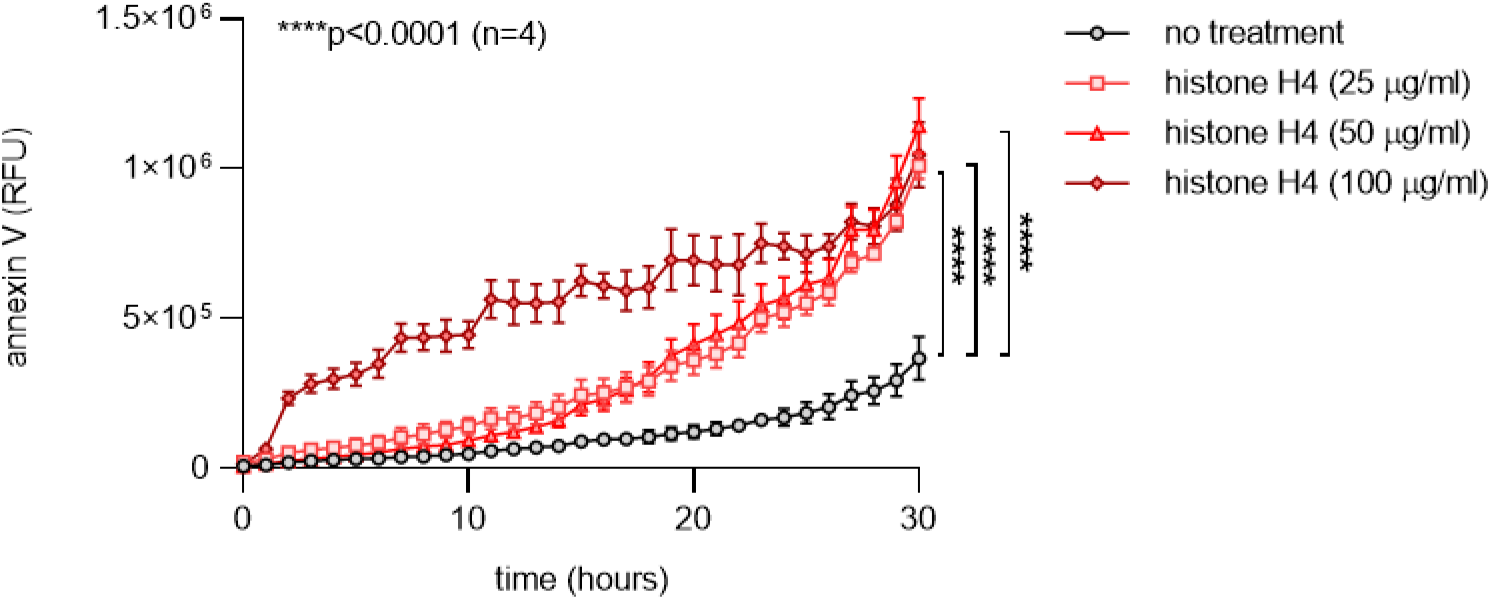
HUVECs expose phosphatidylserine in response to histone H4. HUVECs were treated with different concentrations of histone H4 in the presence of Annexin V red agent. The plate was imaged every hour using the IncuCyte^®^ S3 timelapse microscope for 30 hours. Mean ± standard deviation for three independent experiments is presented; ****p<0.0001by two-way ANOVA corrected by Dunnett’s test.

**Supplementary Figure 11:**
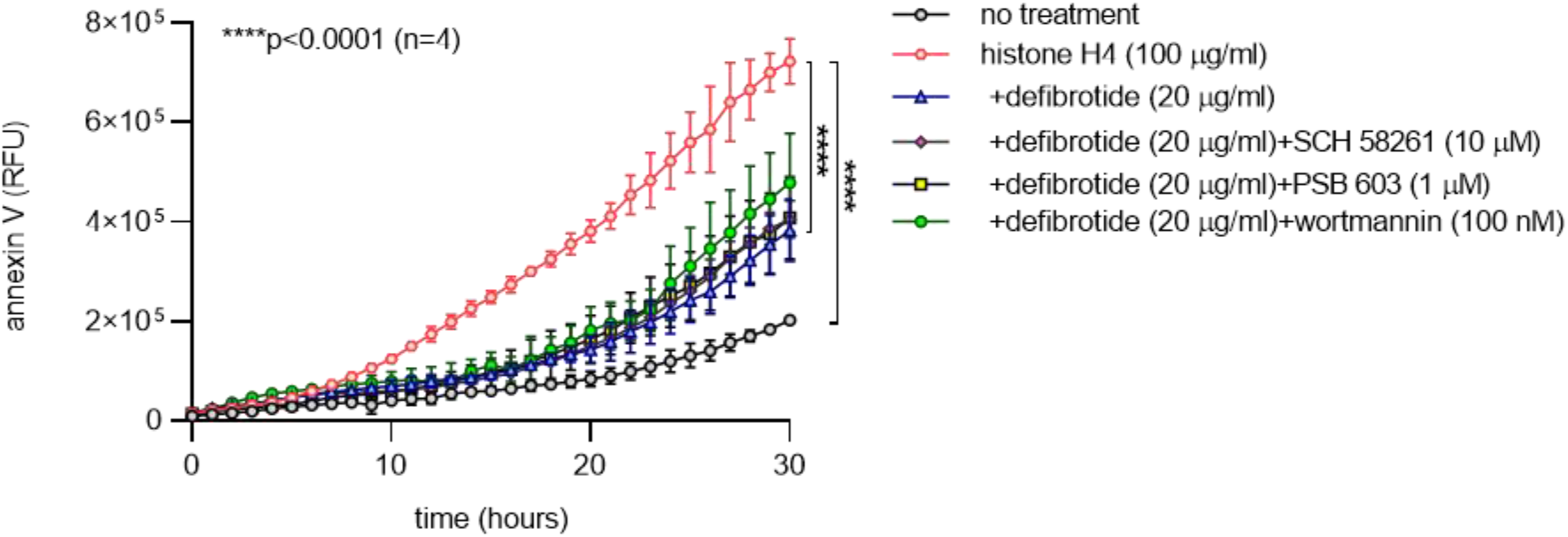
Adenosine receptor antagonists and wortmannin do not abolish the protective effect of defibrotide on HUVECs exposing phosphatidylserine in response to histone H4. HUVECs were treated with histone H4 and defibrotide as indicated. Some samples were additionally treated with the adenosine A_2A_ receptor antagonist SCH 58261, the adenosine A_2B_ receptor antagonist PSB 603, or wortmannin in the presence of Annexin V red agent. The plate was imaged every hour using the IncuCyte^®^ S3 timelapse microscope for 30 hours. Mean ± standard deviation for three independent experiments is presented; ****p<0.0001by two-way ANOVA corrected by Dunnett’s test.

**Supplementary Figure 12:**
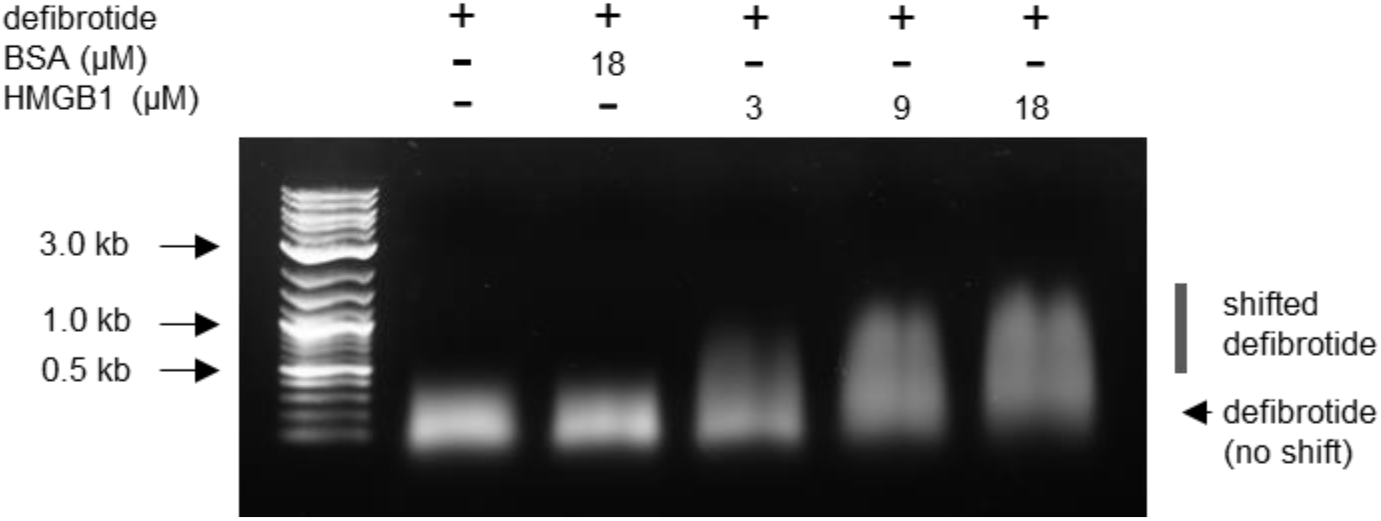
Direct interaction between HMGB1 and defibrotide revealed by electrophoretic mobility shift assay. Defibrotide and HMGB1 were incubated at 37°C for 30 minutes and then resolved on a 0.5% agarose gel.

**Supplementary Figure 13:**
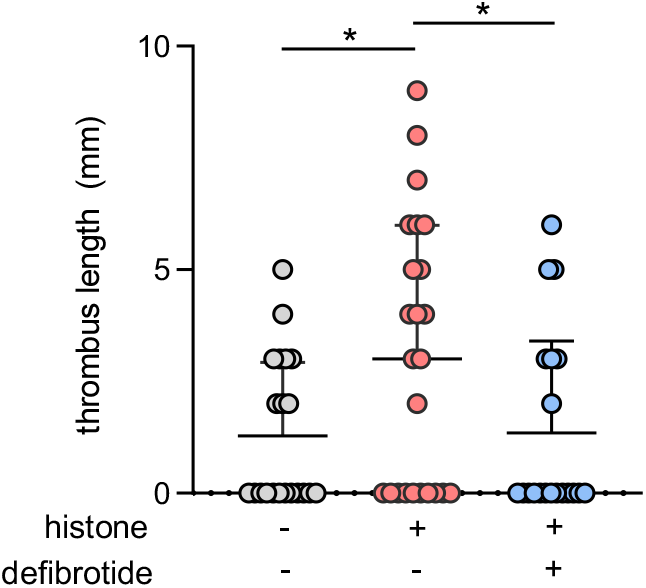
Defibrotide reduces the length of histone-mediated venous thrombi. Mice were injected with either histone (10 mg/kg) or saline via tail vein 1 hour prior to surgery via tail. Meanwhile, defibrotide (150 mg/kg) or saline was administered by retro-orbital injection 24 hours prior to surgery and then immediately following closure of the abdomen. Thrombus length was determined 24 hours later. Scatter plots are presented with each data point representing a unique mouse (horizontal bars=means; *p<0.05 by one-way ANOVA corrected by Dunnett’s test. Data is presented by mean ± standard deviation.

**Supplementary Figure 14:**
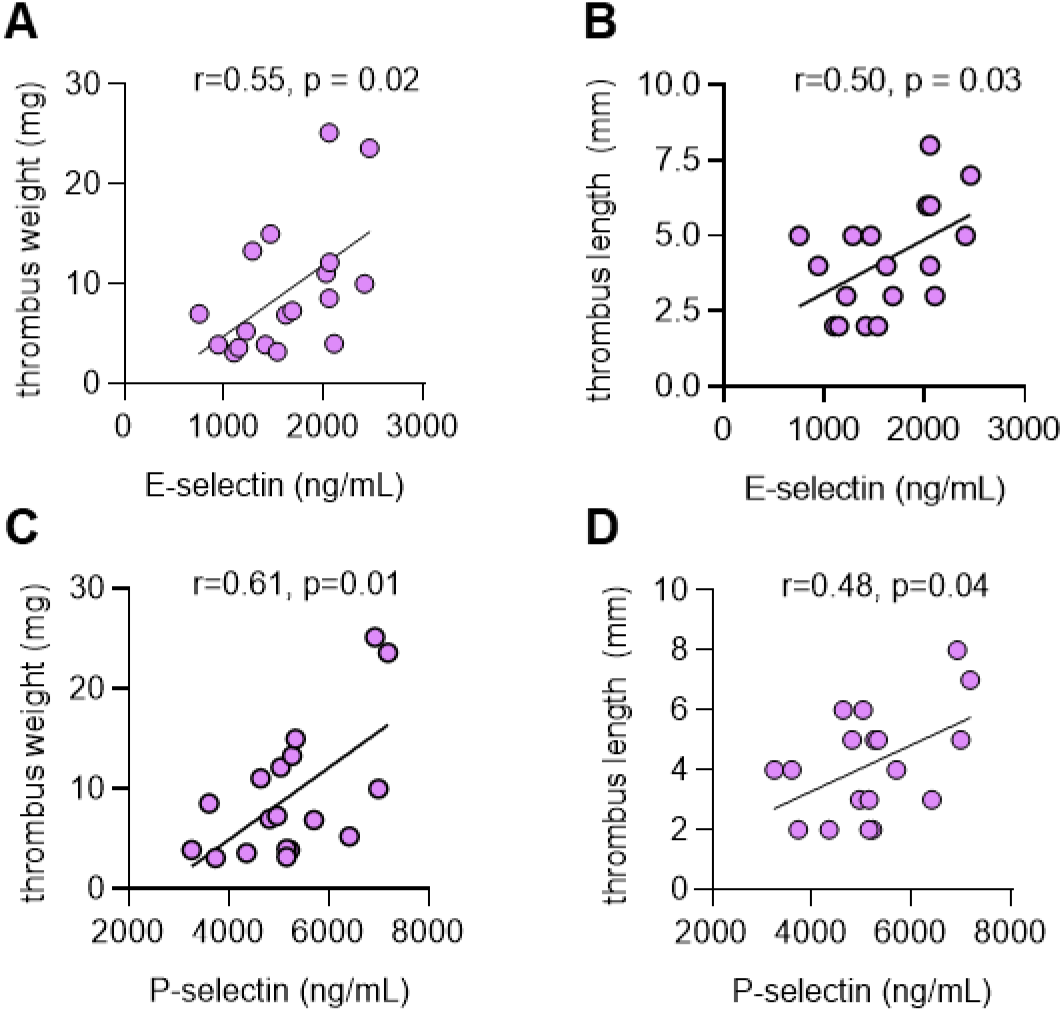
Association between soluble E-selectin or soluble P-selectin and thrombus size in mice. **A-D,** the data presented in Figure 7 and Supplementary Figure 13 are presented here as scatter plots. Correlations were tested by Pearson’s method.

